# Contrast sensitivity: a fundamental limit to vision restoration after V1 damage

**DOI:** 10.1101/2023.08.31.23294827

**Authors:** Jingyi Yang, Elizabeth L. Saionz, Matthew R. Cavanaugh, Berkeley K. Fahrenthold, Michael D. Melnick, Duje Tadin, Farran Briggs, Marisa Carrasco, Krystel R. Huxlin

**Author notes:** Corresponding author: Krystel R. Huxlin, PhD, Flaum Eye Institute, University of Rochester Medical Center, 601 Elmwood Avenue, Box 314, Rochester, NY, 14642, USA.

## Abstract

Stroke damage to the primary visual cortex (V1) causes severe visual deficits, which benefit from perceptual retraining. However, whereas training with high-contrast stimuli can locally restore orientation and direction discrimination abilities at trained locations, it only partially restores luminance contrast sensitivity (CS). Recent work revealed that high-contrast discrimination abilities may be preserved in the blind field of some patients early after stroke. Here, we asked if CS for orientation and direction discrimination is similarly preserved inside the blind field, to what extent, and whether it could benefit from a visual training intervention. Thirteen subacute (<3 months post-V1-stroke) and 12 chronic (>6 months post-V1-stroke) participants were pre-tested, then trained to discriminate either orientation or motion direction of Gabor patches of progressively lower contrasts. At baseline, more subacute than chronic participants could correctly discriminate the orientation of high-contrast Gabors in their blind field, but all failed to perform this task at lower contrasts, even when 10Hz flicker or motion direction were added. Training improved CS in a greater portion of subacute than chronic participants, but no-one attained normal CS, even when stimuli contained flicker or motion. We conclude that, unlike the near-complete training-induced restoration of high-contrast orientation and direction discrimination, there is limited capacity for restoring CS after V1 damage in adulthood. Our results suggest that CS involves different neural substrates and computations than those required for orientation and direction discrimination in V1-damaged visual systems.

**Significance statement:** Stroke-induced V1 damage in adult humans induces a rapid and severe impairment of contrast sensitivity for orientation and direction discrimination in the affected hemifield, although discrimination of high-contrast stimuli can persist for months. Adaptive training with Gabor patches of progressively lower contrasts improves contrast sensitivity for these discriminations in the blind-field of both subacute (<3 months post-stroke) and chronic (>6 months post-stroke) participants, although it fails to restore fully-normal contrast sensitivity. Nonetheless, more subacute than chronic stroke participants benefit from such training, particularly when discriminating the orientation of static, non-flickering targets. Thus, contrast sensitivity appears critically dependent on processing within V1, with perceptual training displaying limited potential to fully restore it after V1 damage.

## Introduction

Damage to the primary visual cortex (V1) causes cortically-induced blindness (CB) – a prominent, homonymous visual impairment through both eyes, which afflicts 25-50% of stroke patients (Hazelton et al., 2019; Zhang et al., 2006). By 6 months post-stroke, CB participants are considered “chronic” and exhibit stable defects in detecting and discriminating targets presented to the hemifield contralateral to their V1 damage (reviewed by Saionz et al., 2022). Notwithstanding the immediate vision loss measured by clinical perimetry (Townend et al., 2007), some CB participants retain either unconscious [i.e., blindsight (reviewed by Weiskrantz, 1996)] or conscious, residual visual abilities (Cavanaugh et al., 2023; Redmond et al., 2023; Riddoch, 1917; Saionz et al., 2020) inside their perimetrically-defined blind fields. Such residual vision differs from normal in many ways, including narrower spatiotemporal bandwidth, with best performance at low spatial and mid-high temporal frequencies (Barbur et al., 1994; Morland et al., 1999; Sahraie et al., 2003, 2008). Discrimination abilities also appear more impacted: except for a few subacute post-stroke individuals (Saionz et al., 2020; Cavanaugh et al., 2023), most CB patients fail to consciously discriminate opposite motion directions or differences in orientation (Cavanaugh et al., 2015, 2019, 2023; Das et al., 2014) or to effectively integrate across motion directions (Cavanaugh & Huxlin, 2017; Das et al., 2014; Huxlin, 2009; Saionz et al., 2020) in their blind-field. In addition, after V1 strokes, spatial and temporal contrast sensitivity (CS) functions for discrimination can no longer be reliably measured inside perimetrically-defined blind fields (Clatworthy et al., 2013; Das et al., 2014; Hess & Pointer, 1989). As an essential attribute of vision (Campbell, 1983; Kelly, 1975), the restoration of CS is thus highly sought-after in visually-impaired populations.

Visual training protocols – usually with high-contrast stimuli – are increasingly used in attempts to recover visual abilities in CB fields. Functions that can be improved with such training include fine and coarse direction discrimination and integration (Cavanaugh et al., 2015, 2019; Das et al., 2014; Huxlin, 2009), simple orientation and direction discrimination (Das et al., 2014), detection of flickering gratings (Sahraie et al., 2006, 2010; Trevethan et al., 2012), relative target localization (Chokron et al., 2008; Elshout et al., 2016), flicker sensitivity (Raninen et al., 2007), motion coherence (Vaina et al., 2014), and letter identification (Chokron et al., 2008; Raninen et al., 2007). Some of these training interventions reduced the size of perimetrically-defined blind fields (Cavanaugh & Huxlin, 2017; Elshout et al., 2016), and improved luminance CS, although in no case did CS return to normal (Das et al., 2014; Huxlin, 2009; Saionz et al., 2020). The fact that several of these training interventions partially recovered CS is encouraging, but their inability to recover normal CS at trained, blind-field locations means that recovered vision remains fundamentally degraded and less useful in everyday life.

Why does CS fail to fully recover at trained, blind-field locations in V1-damaged participants? Possible explanations include: (1) training with high-contrast stimuli, which is typical of most vision restoration interventions, might not be optimal for improving CS; (2) CS recovery may depend on *when* training is administered post-stroke (Saionz et al., 2020); and (3) residual visual circuitry may be permanently altered by the V1 damage, and restoring normal CS may simply not be possible. In intact visual systems, CS correlates positively with V1 surface area (Himmelberg et al., 2022). V1 neurons are highly selective to contrast and underlie contrast discrimination (Albrecht & Hamilton, 1982; Boynton et al., 1999). If these neurons are lost, the residual visual system may simply lack the neural circuitry necessary to fully recover CS, regardless of timing of training and/or training protocol. To arbitrate among these possibilities, we compared the efficacy of training in subacute *versus* chronic CB stroke patients, with stimuli of progressively lower luminance contrasts, at spatial and temporal frequencies optimal for stimulus detection and discrimination in the blind-field (Sahraie et al., 2003, 2008).

## Materials and Methods

### Participants

Thirteen subacute CB participants (mean ± standard deviation (SD): 7.5 ± 3.8 weeks post-stroke) and 12 chronic CB participants (27.6 ± 33.0 months post-stroke) took part in this study. Participant demographics, training assignments, and history are detailed in **Table 1**. Although 6 chronic participants were naïve, three (CB003, CB008, and CB011) trained first as subacutes and then as chronics in the present study. Another set of 3 chronic participants participated in two prior, unrelated studies, where they learned to discriminate the motion of high-contrast, random dot stimuli (Saionz et al., 2020; Cavanaugh et al., 2022) (as detailed in **Table 1**). All participants sustained stroke-induced, unilateral damage to the occipital lobe in adulthood, causing unilateral, homonymous visual field defects (**Fig. 1**). Stroke damage was confirmed by brain imaging (**Fig. 1**), and visual defects were confirmed using 10-2 and 24-2 Humphrey visual fields (HVF), collected and analyzed as described below (Cavanaugh & Huxlin, 2017). Eligible participants were required to have reliable HVFs (fixation losses, false positives, and false negatives <20%); they had to demonstrate stable and accurate fixation verified by an eye-tracker during in-lab psychophysical testing (see below), and they had to be free of ocular or neurological diseases that could interfere with or confound the reason for poor perceptual performance. All participants were best corrected using glasses or contact lenses during testing and training. The Research Subjects Review Board approved study procedures at the University of Rochester, which were conducted as per the Declaration of Helsinki, with written informed consent obtained from each participant, and participation was voluntary.

**Figure 1.**
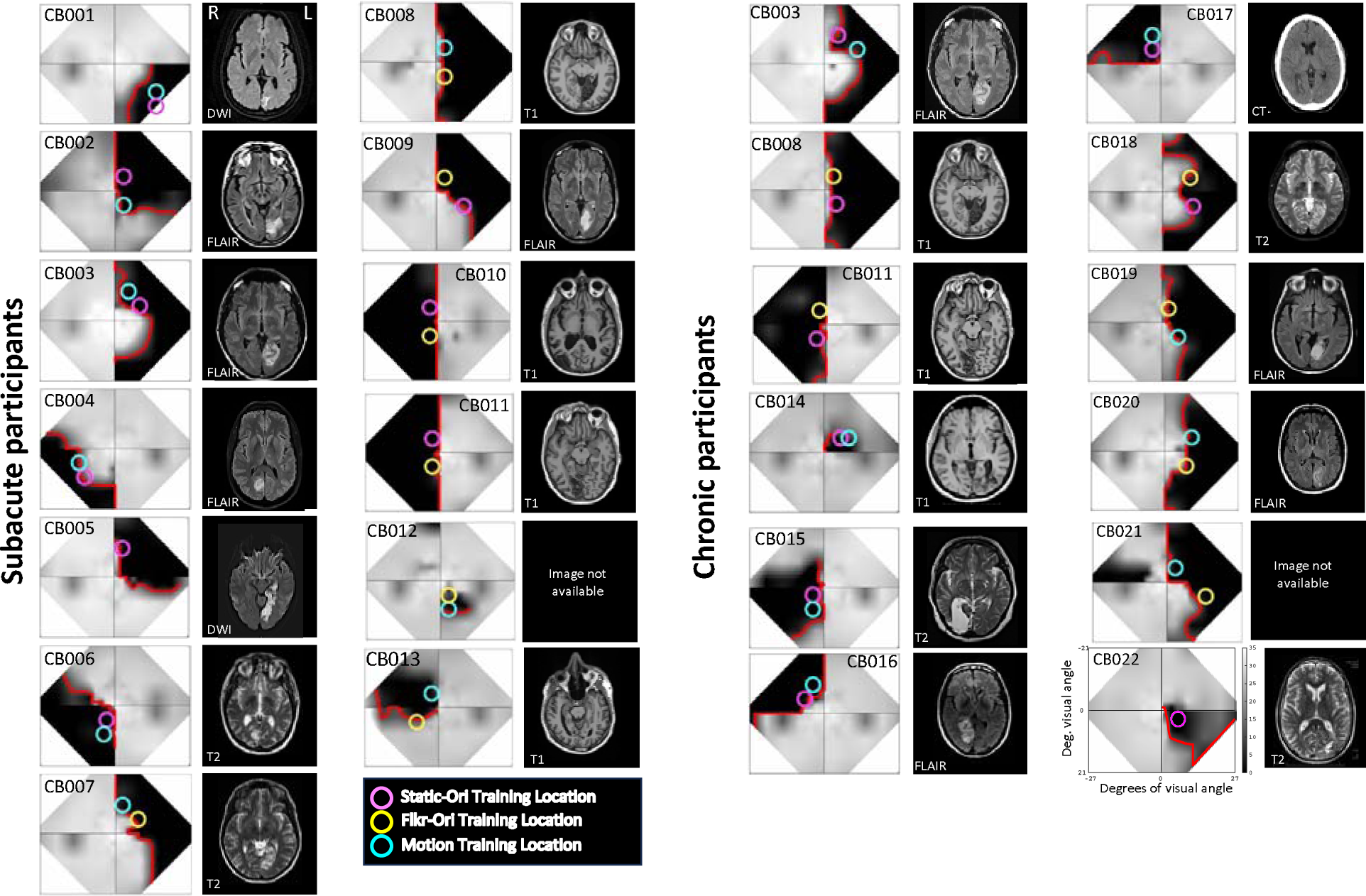
Composite visual field maps, locations selected for training and brain scans for individual CB participants separated into subacute and chronic groups. Humphrey perimetry-derived, binocular maps of visual sensitivity (grey scale next to CB022, dB)a cross the central visual field, alongside sample clinical brain images illustrating stroke damage to the occipital cortex. Brain imaging type [T1, T2, diffusion-weighted imaging (DWI), T2-weighted fluid-attenuated inversion recovery (FLAIR) for magnetic resonance imaging; computed tomography (CT)] are indicated on each radiographic image, which is shown according to radiographic convention with left brain hemisphere (L) on image *right*. In visual field maps, the red line denotes the blind field border encircling regions of mean Humphrey sensitivity <10dB, according to the Social Security Administration definition of blindness (see main text for details). Colored circles on each visual field map represent the locations and size of Gabor stimuli used to train participants on the three different discrimination tasks: Static-Ori (Task 1), Flkr-Ori (Task 2) and Motion (Task 3).

**Table 1.** Participant demographics and training assignments. Task 1: Static-Ori, orientation discrimination of a static, non-flickering Gabor; Task 2: Flkr-Ori, Orientation discrimination of a flickering Gabor; Task 3: Motion, direction discrimination of a drifting Gabor; FDD: fine direction discrimination; DI: direction integration; SA: subacute; CH: chronic.

| Patient code | Sex | Time post-stroke | Training tasks \| Loc. 1, 2 | Prior training (phase) | Prior training task | Pre-post interval (months) | Compliance \| Loc. 1, 2 (%) | Deficit area (deg<sup>2</sup>) | Eccentricity \| Loc. 1, 2 (deg) |
| --- | --- | --- | --- | --- | --- | --- | --- | --- | --- |
| Subacute (SA) participants |  |  |  |  |  |  |  |  |  |
| CB001 | Male | 2.4 wks | Task 1 | No | N/A | 5.4 | 34 | 233 | 21.2 |
| CB002 | Female | 6.7 wks | Task 1, 3 | No | N/A | 4.9 | 126, 128 | 576 | 5.8, 5.8 |
| CB003 | Male | 6 wks | Task 1, 3 | No | N/A | 4.6 | 100, 106 | 586 | 10.3, 11.2 |
| CB004 | Male | 2 wks | Task 1, 3 | No | N/A | 5.6 | 91, 90 | 291 | 14.9, 13.9 |
| CB005 | Female | 8.1 wks | Task 1 | No | N/A | 4.5 | 19 | 451 | 10.4 |
| CB006 | Male | 3.1 wks | Task 1, 3 | No | N/A | 5.6 | 66, 75 | 517 | 5.8, 10.8 |
| CB007 | Female | 5 wks | Task 2, 3 | No | N/A | 5.4 | 93, 92 | 509 | 9.4, 10.3 |
| CB008 | Male | 11.4 wks | Task 2, 3 | No | N/A | 4.0 | 128, 126 | 753 | 5.8, 5.8 |
| CB009 | Male | 14 wks | Task 1, 2 | No | N/A | 3.7 | 126, 129 | 587 | 11.2, 5.8 |
| CB010 | Male | 10.4 wks | Task 1, 2 | No | N/A | 5.0 | 108, 106 | 807 | 5.6, 5.6 |
| CB011 | Female | 11.4 wks | Task 1, 2 | No | N/A | 4.9 | 113, 115 | 804 | 5.6, 5.6 |
| CB012 | Male | 7.7 wks | Task 2, 3 | No | N/A | 5.1 | 85, 85 | 68 | 5.8, 10.8 |
| CB013 | Male | 9.4 wks | Task 2, 3 | No | N/A | 5.3 | 132, 132 | 310 | 9.4, 5.6 |
| **Mean±SD** |  | **7.5±3.8 wks** |  |  |  | **4.9±0.6** | **98±32** | **500±226** | **9.2± 4.3** |
| Chronic (CH) participants |  |  |  |  |  |  |  |  |  |
| CB003 | Male | 9.8 mo | Task 1, 3 | Yes (SA, CH) | Task 1, 3, FDD | 3.1 | 113, 113 | 534 | 11.2, 13 |
| CB008 | Male | 6.6 mo | Task 1, 2 | Yes (SA) | Task 2, 3 | 8.2 | 109, 109 | 740 | 6.4, 5.8 |
| CB011 | Female | 7.7 mo | Task 1, 2 | Yes (SA) | Task 1, 2 | 8.5 | 96, 98 | 783 | 6.1, 5.6 |
| CB014 | Male | 6.9 mo | Task 1, 3 | Yes (SA) | DI | 17.8 | 79, 82 | 65 | 7.8, 10.3 |
| CB015 | Female | 42.8 mo | Task 1, 3 | Yes (CH) | DI, FDD | 5.9 | 47, 47 | 447 | 4.7, 8.5 |
| CB017 | Male | 33.2 mo | Task 1 | Yes (CH) | FDD | 3.5 | 59 | 377 | 5.8 |
| CB016 | Female | 24.7 mo | Task 1, 3 | No | N/A | 4.7 | 118, 125 | 373 | 8.6,10.8 |
| CB018 | Male | 121.4 mo | Task 1, 2 | No | N/A | 5.1 | 64,64 | 554 | 12.1,11.2 |
| CB019 | Female | 48.7 mo | Task 2, 3 | No | N/A | 9.6 | 66, 69 | 646 | 5.6,7.8 |
| CB020 | Female | 6.3 mo | Task 2, 3 | No | N/A | 5.8 | 102, 103 | 576 | 9.4,11.2 |
| CB021 | Female | 7.5 mo | Task 2, 3 | No | N/A | 5.2 | 86, 89 | 726 | 14.9,5.8 |
| CB022 | Male | 15.6 mo | Task 1 | No | N/A | 4.7 | 89 | 167 | 6.7 |
| **Mean±SD** |  | **28±33 mo** |  |  |  | **6.8±4.0** | **86±23** | **499±224** | **8.6± 2.9** |

### Perimetric mapping of visual field defects

Monocular 10-2 and 24-2 HVF perimetry was conducted using a Humphrey Field Analyzer II-i750 system (Zeiss Humphrey Systems, Carl Zeiss Meditec) at the University of Rochester Flaum Eye Institute. All tests were performed by the same ophthalmic technicians, with fixation controlled using the system’s eye tracker and gaze/blind spot automated controls. Visual acuity was corrected to 20/20 using trial lenses. A white size III stimulus and a background luminance of 11.3 cd/m^2^ were used.

The resulting four test patterns were interpolated using a custom MATLAB (Mathworks, United States) script to create a unitary, composite HVF map for each participant. Luminance detection thresholds (dB) from monocular HVFs were first averaged between the two eyes and then combined as previously described (Cavanaugh & Huxlin, 2017). The final composite HVF maps (**Fig. 1**) covered a visual field area of 1,616 deg^2^, stretching a maximum of ± 27° along the x-axis and ± 21° in the y-axis. From these maps, we computed the blind-field borders, drawn as a line encompassing regions of the visual field where visual sensitivity was <10dB (red lines on HVF maps, **Fig. 1**). This threshold was chosen per the definition of legal blindness provided by the Social Security Administration (Medical/Professional Relations Disability Evaluation Under Social Security 2.00 Special Senses and Speech-Adult., 2019). Each in-lab psychophysical test location was then drawn as a circle with a radius of either 2 or 2.5 degrees (matching the stimulus size used) onto its corresponding HVF map (**Figs. 1, 3**) before being categorized by computing the distance in degrees between the blind-field border and each stimulus center (see below for details).

### Apparatus for in-lab psychophysics with eye tracking

Visual discrimination tasks were performed on a MacPro computer with stimuli displayed on a CRT monitor (HP 7217A, 48.5×31.5cm screen size, 1024×640 resolution, 120Hz frame rate, or Dell N993s, 36.5×27cm screen size, 800×600 resolution, 120Hz frame rate). In all cases, monitor luminance was calibrated using a ColorCal II automatic calibration system (Cambridge Research Systems, United Kingdom), and the resulting gamma-fit linearized lookup table was implemented in MATLAB. A viewing distance of 42 cm was ensured using a chin/forehead rest. Eye position was monitored binocularly and continuously using an Eyelink 1000 eye tracker (SR Research Ltd., Canada) with a sampling frequency of 1000 Hz and accuracy within 0.25°. All tasks and training were conducted using MATLAB and Psychtoolbox (Brainard, 1997; Pelli, 1997).

### Experimental Design

#### Stimuli and tasks (Fig. 2)

Each participant had two in-lab visits, during which they were tested with a battery of two-alternative, forced-choice (2AFC) discrimination tasks. The first visit was used to assess baseline performance in the blind and intact fields and to select appropriate training locations in the blind field. Patients then performed several months of at-home training on one of the pre-tested tasks, before returning to the laboratory for a repeat of baseline tests (see **Table 1** for testing intervals). This return visit allowed us to measure changes in performance with eye-tracker-enforced fixation at initially-tested locations.

**Figure 2.**
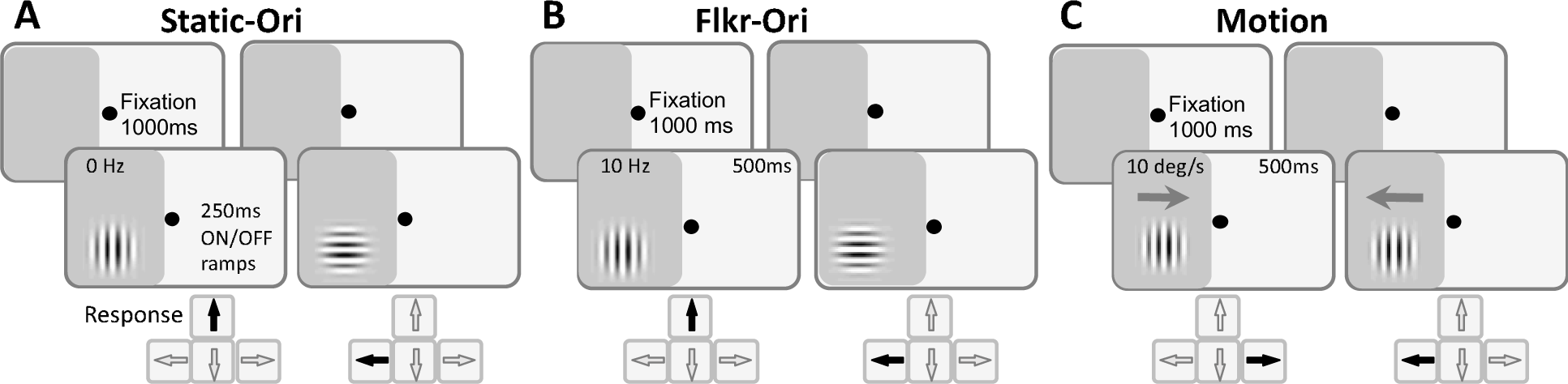
Trial sequences for psychophysical tasks. **A.** Static-Ori (Task 1): vertical *versus* horizontal orientation discrimination of non-flickering, static Gabors that appeared over a 250ms onset, followed by a 250ms offset. **B.** Flkr-Ori (Task 2): vertical *versus* horizontal orientation discrimination of 10Hz-flickering Gabors with a 500ms duration. **C**. Motion (Task 3): left *versus* right direction discrimination of 10 deg/sec drifting Gabors. Participants were asked to first fixate a spot in the middle of the screen for 1000ms for stimuli to appear, illustrated here in the impaired hemifield of vision (dark grey shading). Responses were required after each trial. They involved pressing the indicated arrow key on the computer keyboard, followed by auditory feedback as to the correctness of the response.

Stimuli used for in-lab tests and home training were identical. They consisted of Gabor patches (sinusoidal gratings 4-5° in diameter with a Gaussian envelope, sigma 1°) presented on a uniform, mid-grey background, with a 250ms on/off temporal raised cosine envelope for Task 1, or for 500ms for Tasks 2 and 3. **Task 1 (Fig. 2A)** – vertical *versus* horizontal orientation discrimination of a static, non-flickering Gabor (Static-Ori); **Task 2 (Fig. 2B)** – vertical *versus* horizontal orientation discrimination of a 10Hz flickering Gabor (Flkr-Ori); **Task 3 (Fig. 2C)** – leftward *versus* rightward direction discrimination of a vertical, drifting Gabor (Motion), whose temporal frequency was set to 10Hz, generating a drift speed of 10deg/s. Spatial frequency (SF) was set to 1 cycle per degree (cpd) for all tasks. Within each test set of 100 trials, achromatic luminance contrast was titrated using a single 2-down 1-up staircase with the following steps: 100, 75, 50, 25, 20, 15, 10, 5, 2, 1%. Contrast thresholds were calculated by fitting a Weibull function to 72.5% percent correct performance, selected because it lies halfway between chance (50% correct) and 95% correct performance, assuming a 5% lapse rate. Contrast sensitivity was calculated as the reciprocal of the contrast threshold.

During in-lab testing, contrast sensitivity functions (CSF) (specifically, CS *versus* spatial frequency functions) were also measured for all three tasks using a Bayesian, adaptive, quick (qCSF) method (Hou et al., 2010; Lesmes et al., 2010). The CSF indexes the window of visibility. This approach allowed us to estimate contrast thresholds (72.5% correct) over a broad spatial frequency range in 100 trials (Lesmes et al., 2010). When relevant (i.e., for flickering or drifting Gabors), the temporal frequency was set at 10Hz. For qCSF measurements, stimulus contrast varied between 0.1 and 99% in steps of 1.5 dB, and SF varied from 0.25 to 7.5 cpd. The qCSF was expressed as a truncated log-parabola defined by four parameters: peak sensitivity, peak spatial frequency, bandwidth at half-height, and low-frequency truncation level. This function allows estimation of the area under the curve (a summary of the entire range of contrast visibility) and the high-cutoff SF values (the SF where the contrast threshold is 100%). Participants performed one qCSF run per condition (Static-Ori/Flkr-Ori/Motion), consisting of 100 trials at each visual field location of interest.

#### Mapping and selection of training locations (Fig. 3)

To characterize visual discrimination performance at different contrasts in each participant, we measured performance on the *Static-Ori* task in the intact visual hemifield across the HVF-defined blind-field border and inside the blind field. Mapping usually started in the intact field or straddling the intact/blind-field border (**Fig. 3A, B**), where participants attained relatively normal performance levels (compared to a location of similar eccentricity in their intact hemifield of vision). Each discrimination task began with participants asked to fixate on a centrally presented black spot on the computer monitor in front of them for 1000ms. A visual stimulus appeared (see below for description), accompanied by a tone (especially important to signal stimulus appearance in the blind-field). The eye tracker monitored eye movements within a 2×2° square window centered on the fixation spot. Trials in which eye movements or drifts broke the fixation window were signaled by a loud tone, eliminated from the analysis, and reshuffled into the trial sequence. Participants responded by pressing arrow keys on the computer keyboard, followed by auditory feedback as to the correctness of their response (high-pitched tone = correct response; low-pitched tone = incorrect response). We verified that each participant could hear differences between tones and interpret them correctly. New trials were initiated automatically 500ms after a response or fixation break.

**Figure 3.**
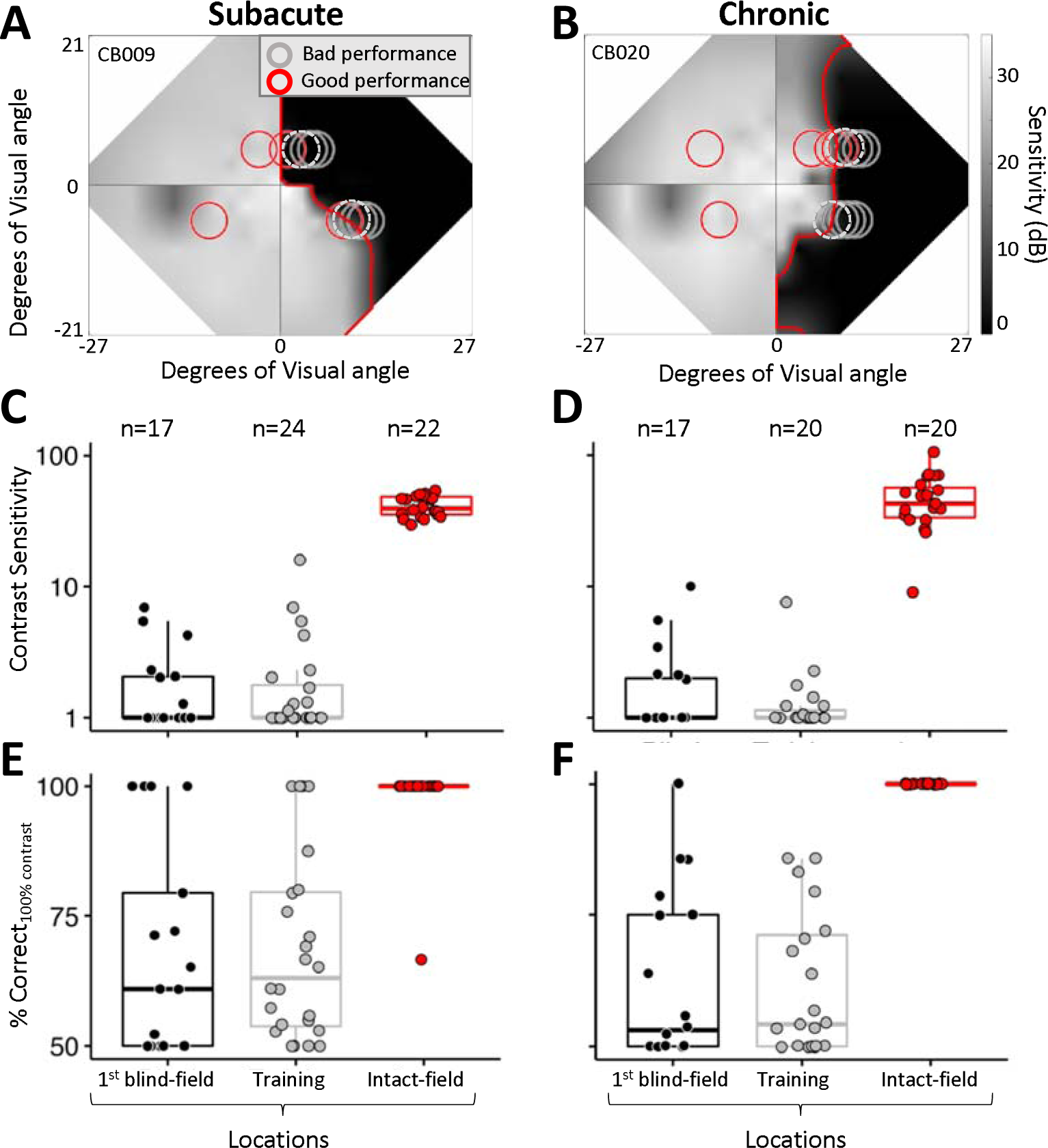
Mapping of baseline performance across the blind field border. **A.** Example of contrast sensitivity (CS) measured at overlapping locations across the blind field border (red line) in a subacute participant (CB009). Stimulus locations (circles) where psychophysical performance was measured in-lab are superimposed on the baseline Humphrey composite map, with greyscale denoting luminance detection sensitivity (dB). Red circles indicate stimulus locations with Static-Ori performance ≥72.5% correct; grey circles denote locations where performance was <72.5% correct; dashed grey circles denote locations selected for training. Mapping across the border of the blind field moved horizontally from intact to deeper blind field locations. Once a training location was selected, performance was also measured at mirror-symmetric, intact-field locations (red circles in left hemifield). **B.** Example of CS measurements across the blind field border in a chronic participant (CB020). Labelling conventions as in A. **C.** Plot of CS for Static-Ori in subacute participants at first locations inside the blind-field border (1^st^ blind-field), selected training locations (Training) and corresponding, intact-field locations. Each data point represents a single location. Box plots indicate the median (line within the box), 25-75% quartile range (box), and 10%-90% range (whiskers). Five SA participants had measurable CS (i.e., CS>1) at 7 different 1st blind-field locations, averaging a CS of 3.5±2.1 at these locations. **D.** Plot of baseline CS in chronic participants at first locations inside the blind field border, selected training locations and corresponding, intact-field locations. Each data point represents a single location. Five CH participants had measurable CS at 6 different 1 blind-field locations, with an average CS of 4.2±3.2 at these locations. Labeling convention as in C. **E.** Percent-correct performance on the Static-Ori task for 100% contrast stimuli measured in subacute participants. Four of the 13 subacute participants attained >72.5% correct at 7 different 1st-blind field locations. Labeling conventions as in C-D. **F.** Percent correct performance at 100% contrast on the Static-Ori task in chronic participants. Five of the 12 chronic participants attained >72.5% correct at 6 different first-blind field locations. Labeling conventions as in C-D.

Participants performed 100 trials of the 1cpd *Static-Ori* task at each mapping location. Performance ≥72.5% correct, with a contrast threshold in the normal range, was “good performance” (red circles, **Fig. 3A, B**), and the stimulus was moved laterally deeper into the blind field by 1 degree. Performance was measured at this new location for another 100 trials. The process was repeated until performance fell to a level that prevented a contrast threshold from being computed (i.e., performance <72.5% correct). In most participants, several additional locations were also tested deeper in the blind field (grey circles, **Fig. 3A, B**) to verify this was a reliable failure point. This mapping procedure allowed us to ascertain how performance changed when stimuli first became fully enclosed within the HVF-defined blind-field border (red line, **Fig. 3A, B**) and to select training locations in each participant.

The locations selected for training (dotted grey circles, **Fig. 3A, B**) were chosen as the first sites where CS fell below that at corresponding locations in the same person’s intact field over a single, 100-trial block after a 1° lateral movement from the intact towards the blind-field. The lowest intact-field CS for subacutes and chronics was 29.6 and 9, respectively. With few exceptions (e.g., CB014, whose deficit was too small, CB005, whose training program did not work correctly at one location, and CB022, who had preserved motion perception across their entire blind field), participants were trained on two different tasks at non-overlapping locations along the vertical meridian (**Fig. 1**, **Fig. 3A, B**). However, it is important to note that training locations were not always fully enclosed within the perimetrically-defined blind-field border; as illustrated by the positioning of some of the dotted grey circles in **Fig. 3A, B**, participants sometimes exhibited abnormal CS at stimulus locations that partially overlapped the intact field. Nonetheless, 24/46 training locations were located entirely in the blind-field, sometimes many degrees deeper than the border. Because of this variance across training locations relative to the border, we compared baseline performance at visual field locations chosen for training and those which first fell entirely within the HVF-defined blind-field border in each participant (**Fig. 3C-F**). Finally, we should also note that while training locations were pre- and post-tested on all three discrimination tasks, each location was trained using only a single task (either Fkr-Ori, Static-Ori, or Motion – see **Fig. 1** for color-coded representation of task assignment per location).

#### At-home visual training

After baseline measurements in-lab, participants were sent home to train for several months (subacute (SA): 4.9±0.6 months [mean ± SD]; chronic (CH): 6.8±4.0 months, **Table 1**). They used their personal computers with a chin/forehead-rest provided by the lab, which they were instructed to position 42cm away from their display during training. Participants performed 300 trials of their assigned training tasks (see **Table 1**, **Fig. 1**) per location per day, at least five days per week, and they emailed their data log files back to the lab for analysis every week. During home training sessions, they were instructed to stay fixated on the fixation spot and warned that inadequate fixation accuracy could limit recovery.

Session thresholds were calculated by fitting a Weibull function with a 72.5% correct performance threshold criterion. After participants’ thresholds improved consistently for at least 20 sessions, we moved their training stimulus 1° deeper into the blind field along the x-axis (Cartesian coordinate space). Once participants trained for ∼4 months, with at least one improved location (defined as consistently good contrast thresholds at that location), they were brought back to the lab, and performance at all home-trained locations was verified with eye tracker-enforced fixation control. We aimed for a similar number of training sessions at the blind-field locations of interest before scheduling people to return for in-lab performance verification. However, the amount of time elapsed until the return visit did vary, as it was affected by the individual’s rate of improvement, their work/family schedules, and ability to travel to our single study site (participants originated from across the entire United States and Canada). For this reason, we also computed participant compliance (**Table 1**), defined as the number of training sessions performed at each blind-field location as a percentage of the number of prescribed training sessions (one training session per location per day multiplied by the number of prescribed training days (5 days/week) between the last day of the pre-training visit and the first day of the post-training visit).

### Statistical analyses

Training locations were treated as independent due to the non-uniform nature of hemianopic visual field defects, both in terms of baseline discrimination performance and training-induced changes in performance (Das et al., 2014; Huxlin et al., 2009; Saionz et al., 2020). Although two locations were sampled in each person’s blind field, only one intact-field location was tested in some participants due to time constraints or participant exhaustion. For this reason, Mann-Whitney (two groups) or Kruskal-Wallis tests with post-hoc Dunn’s multiple comparison tests (more than two groups) were used.

Because of the adaptive nature of the qCSF procedure, a bootstrap method was used to determine statistically significant changes across individuals. Individual trials were first randomly resampled 2,000 times with replacement to generate a resampled trial sequence, which was refitted using the qCSF procedure. From these 2,000 qCSFs at each test location, we repeated the resampling procedure 10,000 times to generate bootstrap distributions of the means of the fitted parameters and the CS means at each SF. We used a peak CS <2.55 as our chance level performance, and thus any trial with peak sensitivity <2.55 was set to zero (for detailed methods, see Saionz et al., 2020). We used a three-way analysis of variance (ANOVAs) for unpaired samples between pre-training and intact-field CSFs. To compute p-values for comparisons between pre- and post-training CSFs, we used paired student t-tests to compare the means of each parameter, and CS means at each SF. For group comparisons at each of the 12 SF levels, p-values were Bonferroni-corrected for multiple comparisons.

## Results

### Baseline CS for static orientation discrimination at 1cpd

*In their intact hemifield of vision*, SA participants, who were all naïve to training, averaged (±SD) 75.2±1.3% correct overall and a CS of 41.7±7.4 on the Static-Ori task; whereas CH participants performed comparably, averaging 75±2% correct with a CS of 48.5±21.5. Individual data for both groups, along with medians and quartiles are shown in **Fig. 3C** (SA) and **Fig. 3D** (CH). Eccentricities tested ranged from 5.6 - 21.2° for SA and from 5.6 - 14.9° for CH participants (**Table 1**), with no effect of eccentricity on CS (SA: r_20_= −0.25, p=0.26; CH: r_18_= -.21, p=0.38).

*At selected training locations*, baseline performance on the Static-Ori task was markedly lower in both SA (61±9% correct, CS=2.3±3.3, **Fig. 3C**) and CH participants (60±9% correct, CS=1.5±1.5, **Fig. 3D**). As with the intact-field locations, eccentricity did not appear to influence CS in either patient group (SA: r_22_= -.06, p=0.78; CH: r_18_=.08, p=0.74). At *first locations inside the perimetrically-defined blind-field border,* SA **(Fig. 3C)** and CH **(Fig. 3D)** participants’ average CS was 2.0±1.8 and 2.1±2.4, respectively. Less than half of participants [SA (5/13), CH (5/12)] had partial preservation of CS (>1), with most of them able to correctly discriminate static orientation at >72.5% correct when stimuli were 100% contrast **(Fig. 3E, F)**.

There was a significant main effect of test location on CS (Kruskal-Wallis, H_2_=86.2, p<0.001), % correct (H_2_=79.2, p<0.001) and % correct at 100% contrast: (H_2_=66.6, p<0.001). A post-hoc Dunn’s test for multiple comparisons using a Bonferroni-adjusted alpha level of 0.017 (0.05/3) showed the main effect of test location to be driven by intact-field locations, where performances were significantly better than at training locations and first locations fully in the blind field. However, there was no significant effect of participant type on performance [CS (H_1_=0.03, p=0.87), % correct (H_1_=0.01, p=0.92), % correct at 100% contrast (H_1_=1.07, p=0.3)]. In sum, the baseline performance of SA and CH participants was comparably good in their intact fields and impaired at first locations fully inside the blind-field border, which were as impaired as locations selected for training. As such, SA and CH participants were well-matched in terms of performance prior to the onset of training. From here on, we will only detail performance changes for selected training locations.

Finally, we should also note that for CH participants, there was no significant impact of prior training on CS (Kruskal-Wallis, H_1_=2, p=0.16) and overall % correct (H_1_=0.16, p=0.69) at all locations tested at baseline. Naïve and previously trained CH participants had similar baseline performance at selected training locations, and at first locations fully inside their blind field.

### Baseline CS for discrimination of flickering and moving Gabors

Key questions in the present study were whether the addition of temporal modulation or motion signals would improve baseline CS in the blind field of people with V1 damage. To answer the first question, we re-measured baseline performance after adding a 10Hz flicker to the orientation discrimination task (Flkr-Ori) or changing the task to a direction discrimination (Motion) at locations selected for training (**Fig. 4**). Kruskal-Wallis tests showed no significant main effects of participant group or task on baseline CS (group: H_1_=0.05, p=0.82; task: H_2_=3.91, p=0.14), overall % correct (group: H_1_=0.41, p = 0.52; task: H_2_=5.12, p = 0.08) or % correct at 100% contrast (group: H_1_=0.08, p=0.77; task: H_2_=4.86, p=0.09). Thus, there was no benefit of adding 10Hz temporal frequency modulation or motion information at blind-field locations selected for training in either SA (**Fig. 4A, C**) or CH (**Fig. 4B, D**) participants.

**Figure 4.**
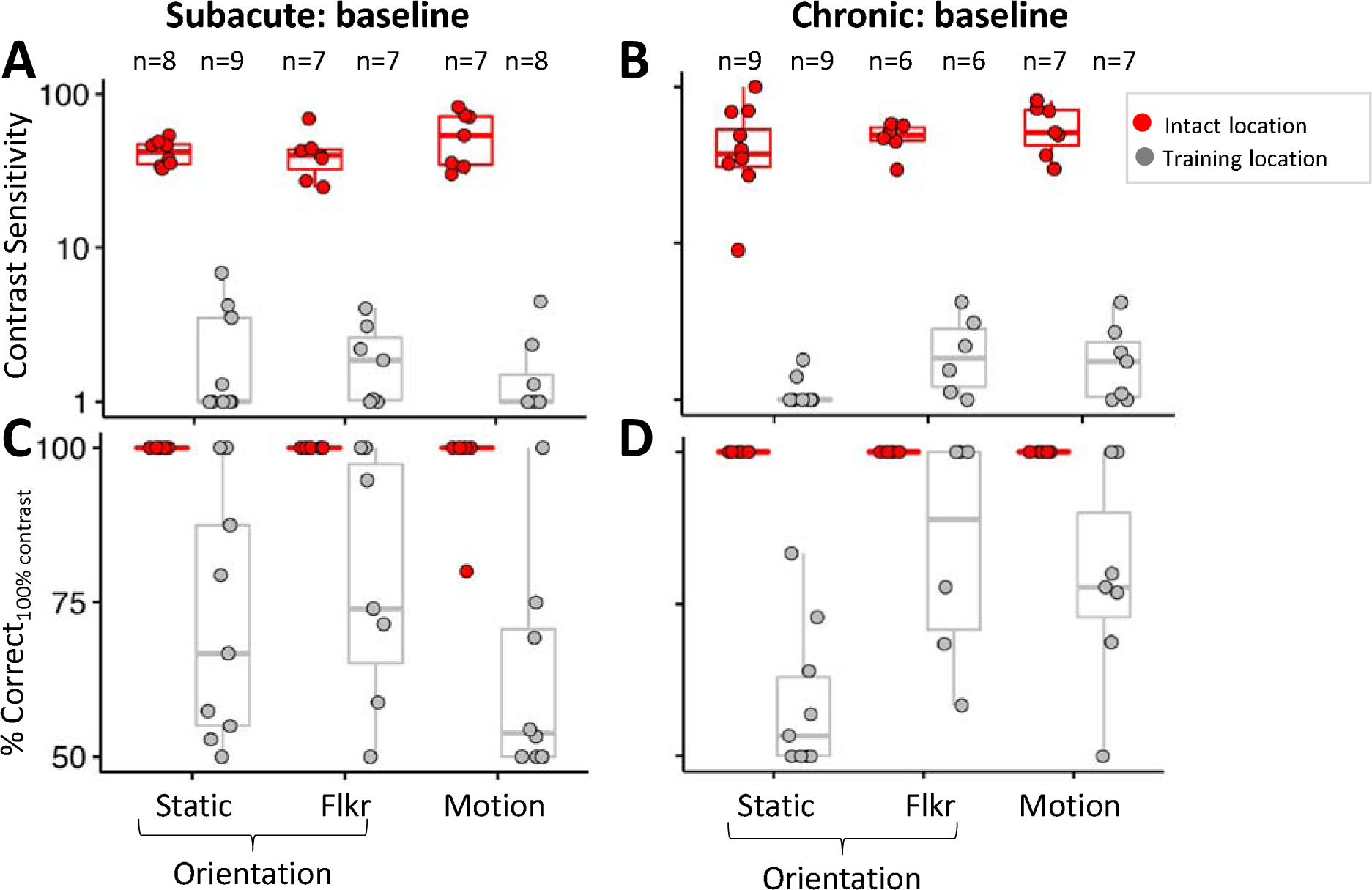
Baseline performance on discrimination of flickering and moving Gabors. **A.** Plot of baseline contrast sensitivity (CS) for orientation discrimination of static Gabors (Static-Ori), flickering Gabors (Flkr-Ori) and direction discrimination of drifting Gabors (Motion) in subacutes at selected training locations and corresponding, intact-field locations. Each data point represents a single location. Red-filled circles indicate performance at corresponding intact-field locations; grey-filled circles denote locations selected for training. Box plots indicate the median (line within the box), 25-75% quartile range (box), and 10%-90% range (whiskers). **B.** Plot of baseline CS for Static-Ori, Flkr-Ori and Motion tasks in chronic participants at selected training locations and corresponding, intact-field locations. Labeling conventions as in A. **C.** Baseline percent-correct performance on the Static-Ori, Flkr-Ori and Motion tasks for 100% contrast stimuli measured in subacute participants. Labeling conventions as in A-B. **D.** Baseline percent-correct performance on the Static-Ori, Flkr-Ori and Motion tasks for 100% contrast stimuli measured in chronic participants. Labeling conventions as in A-C.

### Effect of training on the trained tasks

#### SA participants

Direction/orientation discrimination training improved CS by at least 1 unit in 12/13 SA participants (92%) at 17/24 training locations (71%). Five participants improved at both of their trained locations, 7 at 1 trained location, and 1 at neither trained location. Across tasks, CS improved at 6/9 Static-Ori-trained locations (67%, **Fig. 5A**), 5/7 Flkr-Ori-trained locations (71%, **Fig. 5B**), and 5/8 Motion-trained locations (62.5%, **Fig. 5C**). Overall, post-training CS (mean 8.5±10.2 for three tasks) was significantly higher than pre-training CS (2.0±1.6; Wilcoxon signed-rank, V=205, p=0.0002), although there were no significant differences in CS improvements among training tasks (Kruskal-Wallis, H_2_=0.56, p=0.76). However, post-training CS across all training tasks remained significantly impaired compared to corresponding intact-field locations (45.5±15.8; Wilcoxon rank sum, W=16, p<0.001; **Fig. 7A)**. There was also a significant overall improvement in % correct performance for 100% stimulus contrast (87±18%; Wilcoxon signed-rank, V=183.5, p=0.004), again, without significant differences among training tasks (H_2_=2.87, p=0.24; **Figs. 5D-F**). Post-training overall % correct performance (Wilcoxon rank sum, W=95, p=0.0002) and % correct performance for 100% stimulus contrast (W=172.5, p=0.006) also remained significantly impaired compared to intact-field locations.

**Figure 5.**
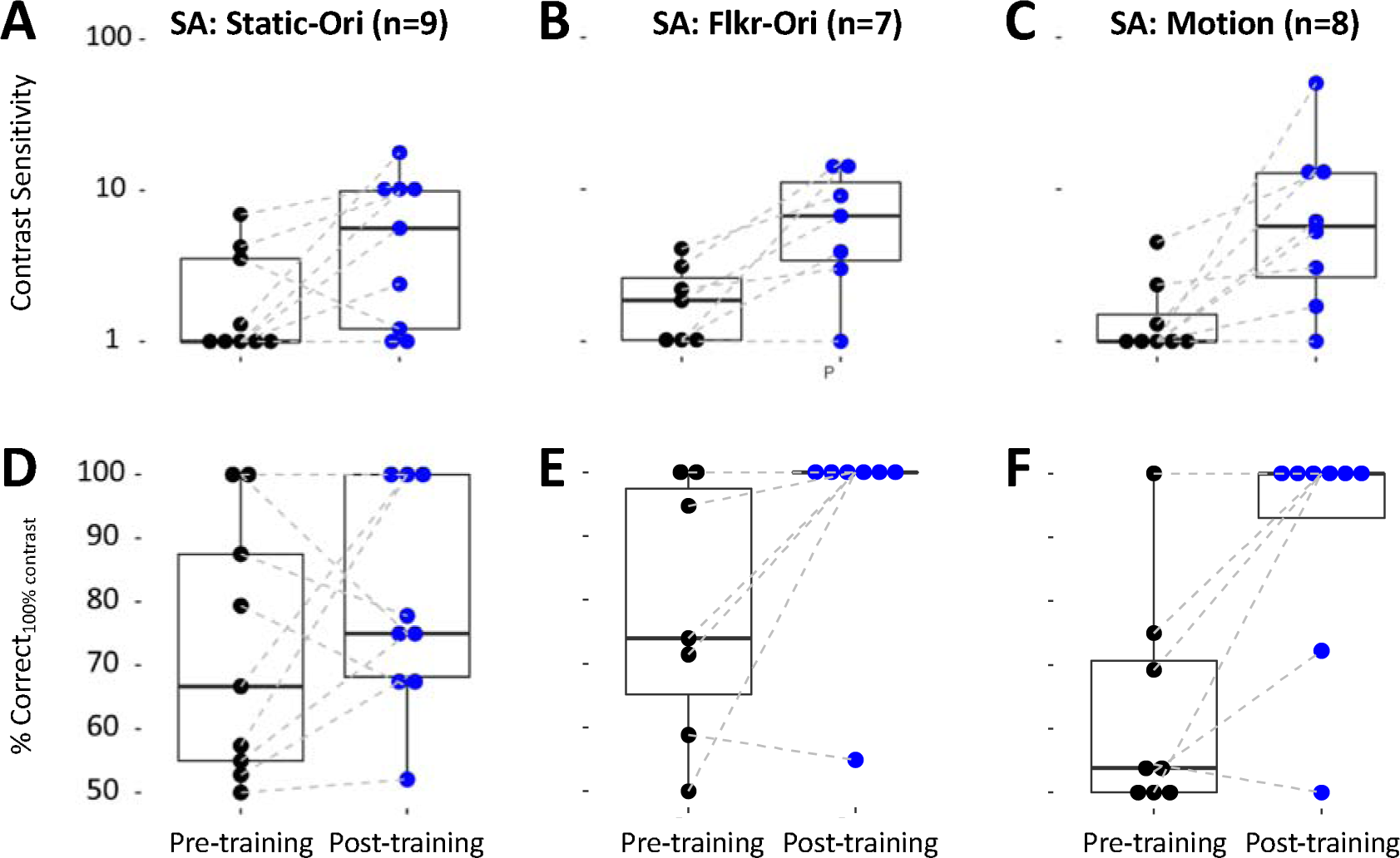
Effect of training on Static-Ori, Flkr-Ori and Motion tasks in subacute participants. **A.** Pre- and post-training contrast sensitivity (CS) for orientation discrimination of static Gabors (Static-Ori) at selected training locations. Each data point represents a single location, with dashed lines connecting the same locations pre- and post-training. Box plots show the median (line within the box), 25-75% quartile range (box), and 10%-90% range (whisker) at each timepoint. **B.** Pre- and post-training CS for orientation discrimination with flickering Gabors (Flkr-Ori) at selected training locations. Labeling conventions as in A. **C.** Pre- and post-training CS for direction discrimination (Motion) at selected training locations. Labeling conventions as in panels A-B. **D.** Pre- and post-training percent-correct performance for 100%-contrast on the Static-Ori task measured at selected training locations. Labeling conventions as in A-C. **E.** Pre- and post-training percent-correct performance on the Flkr-Ori task at 100% contrast, measured at selected training locations. Labeling conventions as in A-D. **F.** Pre- and post-training percent-correct performance on the Motion task at 100% contrast, measured at selected training locations. Labeling conventions as in A-E.

Even when the analysis was restricted to locations that improved, the average CS increase across tasks was 9.8±11.3, with no significant effect of the training task (Kruskal-Wallis, H_2_=1.02, p=0.6). Mean post-training CS was 12±11, which was about six times better than baseline (2.2±1.8; Wilcoxon signed-rank, V=136, p<0.001), but remained about four times lower than mean CS in the same participants’ intact fields (45.2±16.1; Wilcoxon rank sum, W=12, p<0.001).

#### CH participants

Training improved CS by at least 1 unit in fewer people (7/12, 58%) and at fewer trained locations (10/22, 45%) than in SA participants. Of the 7 CH participants whose CS improved, 3 did so at both trained locations, and 4 at 1 location. Across tasks, improvement was seen at 4/9 (44%) locations for the Static-Ori task (**Fig. 6A**), 2/6 (33%) locations for the Flkr-Ori task (**Fig. 6B**), and 4/7 (57%) locations for the Motion task (**Fig. 6C**). Overall, post-training CS (6.9±8) was significantly higher than at baseline (1.7±1; Wilcoxon signed-rank, V=146, p=0.001), with no significant differences in CS improvements among training tasks (Kruskal-Wallis, H_2_=0.67, p=0.72). Unlike SA participants, there was no significant improvement in % correct performance for 100% contrast stimuli (Wilcoxon signed-rank, V=145, p=0.14; **Figs. 6D-F**). As such, post-training CS (Wilcoxon rank sum, W=6, p<0.001; **Fig. 7B**), % correct (W=58.5, p<0.001), and % correct for 100% stimulus contrast (W=66, p<0.001) remained significantly impaired in CH participants compared to their intact-field locations.

**Figure 6.**
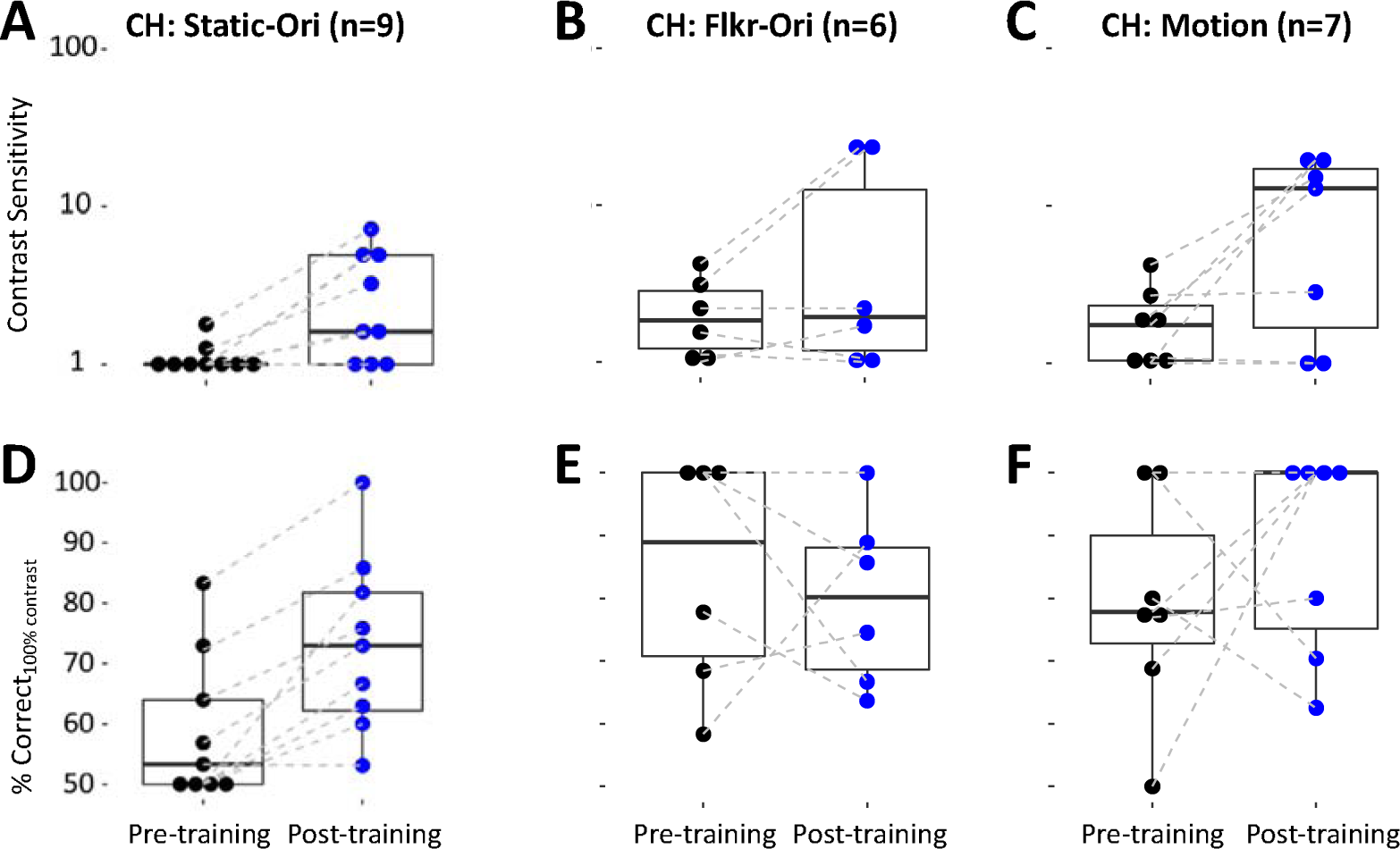
Effect of training on Static-Ori, Flkr-Ori and Motion tasks in chronic participants. **A.** Pre- and post-training contrast sensitivity (CS) for orientation discrimination with static Gabors (Static-Ori) at selected training locations. Each data point represents a single location, with dashed lines connecting the same locations pre- and post-training. Box plots denote the median (line within the box), 25-75% quartile range (box), and 10%-90% range (whiskers). **B.** Pre- and post-training CS for orientation discrimination with flickering Gabors (Flkr-Ori) at selected training locations. Labeling conventions as in A. **C.** Pre- and post-training CS for direction discrimination (Motion) at selected training locations. Labeling conventions as in A-B. **D.** Pre- and post-training percent-correct performance on the Static-Ori task at 100% contrast, measured at selected training locations. Labeling conventions as in A-C. **E.** Pre- and post-training percent-correct performance on the Flkr-Ori task at 100% contrast, measured at selected training locations. Labeling conventions as in A-D. **F.** Pre- and post-training percent-correct performance on the Motion task at 100% contrast, measured at selected training locations. Labeling conventions as in A-E.

When restricting the analysis to the 10 impaired locations that improved in CH participants, the average increase in CS was 11.3±7.2, but the training task had a significant effect (Kruskal-Wallis, H_2_=7.9, p=0.02). A post-hoc Dunn’s test for multiple comparisons using a Bonferroni-adjusted alpha level of 0.017 (0.05/3) showed this effect to be driven by a smaller CS change at Static-Ori-trained locations (3.8±1.6) than at Flkr-Ori- and Motion-trained locations. Finally, mean post-training CS at improved locations across all 3 tasks reached 13.4±8, about 6 times better than the baseline (2.1±1.3, Wilcoxon signed-rank, V=55, p=0.002), but once again, about 4 times less than CS in the same participants’ intact fields (59.2±21, Wilcoxon rank sum: W=0, p<0.001).

In summary, most SA participants showed CS improvement following training, and these improvements were similar across tasks. Conversely, only half of CH participants had improved CS, and for them, adding temporal frequency content generated greater improvements. However, in all cases (SA and CH), post-training CS remained significantly impaired relative to that in the intact fields (**Fig. 7**). Even when restricting the analysis to blind-field locations that benefited from training, CS remained ∼4-5 times lower than in the intact fields, with no significant differences in CS improvements between participant type (SA *or* CH, Kruskal-Wallis, H_1_=0.8, p=0.37). However, the training task had a significant effect on overall CS changes at improved locations (H_2_=6.58, p=0.04). A post-hoc Dunn’s test for multiple comparisons showed this effect to be driven by a smaller CS change at Static-Ori-trained locations (5.6±4.4) than at Motion-trained locations (15.2±13.6). Only one SA Motion-trained participant attained a CS of 50 (asterisk in **Fig. 6A**), which fell within the intact-field range for this task (CS=30-82).

**Figure 7.**
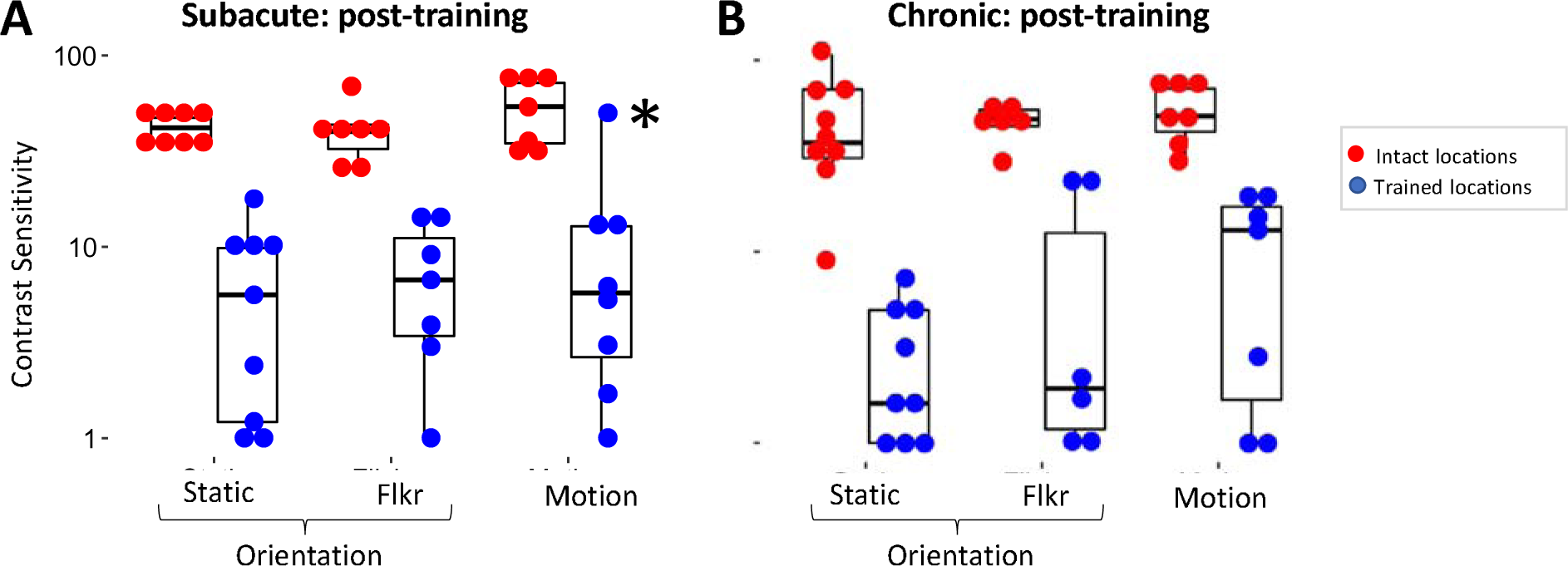
Post-training contrast sensitivity rarely attains intact-field levels. **A.** Contrast sensitivity (CS) for orientation discrimination of static Gabors (Static-Ori), flickering Gabors (Flkr-Ori) and direction discrimination of drifting Gabors (Motion) in subacute participants at trained, blind-field (blue) and corresponding, intact-field (red) locations. Each data point represents a single location. Box plots indicate the median (line within the box), 25-75% quartile range (box), and 10%-90% range (whiskers). Post-training CS at blind-field locations is significantly lower than at corresponding, intact-field locations (Wilcoxon rank sum tests - Static-Ori: W=0, p=0.0006; Flicker-Ori: W=0, p=0.0006; Motion: W=3, p=0.002). **B.** Post-training CS for Static-Ori, Flkr-Ori and Motion tasks in chronic participants at trained, blind-field locations (blue) and corresponding, intact-field locations (red). Post-training CS at blind-field locations is significantly lower than at corresponding, intact-field locations (Wilcoxon rank sum - Static-Ori: W=0, p=0.0004; Flicker-Ori: W=0, p=0.002; Motion: W=0, p=0.002).

### Effect of training amount and compliance on CS change

On average, SA participants trained for 49±36, 34±27, and 53±37 sessions of 300 trials each at Static-Ori, Flkr-Ori, and Motion-trained locations, respectively, equating to compliance (or training density) of 87±40%, 112±18%, and 104±22%. CH participants trained for 59±44, 60±24, and 43±22 sessions at Static-Ori, Flkr-Ori, and Motion-trained locations, equating to compliance of 86±25%, 88±19%, and 90±27%, respectively. Thus, compliance was excellent in the present study, and there was no significant effect of participant group (Wilcoxon rank sum, W=355.5, p=0.05) or training task (Kruskal-Wallis, H_2_=1.31, p=0.52) on compliance. Importantly, we found no significant correlations between compliance (SA: r=-.09, p=0.55; CH: r=-.04, p=0.79) or number of training sessions (SA: r=-.14, p=0.33; CH: r=-.16, p=0.34) and changes in CS attained by either SA or CH participants. Thus, it is unlikely that more training would have further improved performance.

### Transfer of learning to untrained spatial frequencies

The qCSF method estimates the CS *versus* SF function as a truncated log-parabola with four parameters. Prior to training, a three-way ANOVA analysis (**Fig. 8A-F**) confirmed severely impaired qCSFs at locations selected for training *versus* intact-field locations, whether in terms of peak spatial frequency (F_(1,77)_=12.64, p=0.0006), bandwidth at half-height (F_(1,77)_=32.12, p<0.001), and low-frequency truncation level (F_(1,77)_=99.29, p<0.001). Unsurprisingly, differences extended to mean area-under-the-curve (AUC) (pre-training loc: 1.5±1.4, intact loc: 72.6±2.6, F_(1,77)_=594.33, p<0.001) and mean peak CS (pre-training loc: 2.3±1.7, intact loc: 58.8±1.7, F_(1,77)_=923.67, p<0.001). There was a significant interaction between location and task (F_(2,77)_=4.42, p=0.02) on peak CS; this was driven by higher intact-field peak CS for motion (99.5±1.6) than Flkr-Ori (45.1±1.5, p=0.002) and Static-Ori (48.1±1.6, p=0.002), together with a lack of such differences at locations selected for training. Motion qCSFs also had a lower mean low-frequency truncation level (reflecting better sensitivity at low SFs; 0.2±1.4) and narrower mean bandwidth (1.7±1.3) at intact field locations compared to Flkr-Ori and Static-Ori qCSFs, and a smaller mean peak SF (0.8±1.5) compared to static-Ori qCSFs at intact field locations. Importantly, these results showed no significant effect of the participant group (SA *versus* CH) on baseline qCSFs in terms of the abovementioned parameters. In sum, qCSFs were comparable in SA and CH participants at baseline, with no consistent benefit of adding temporal frequency modulation or motion information at blind-field locations selected for training in either group.

**Figure 8.**
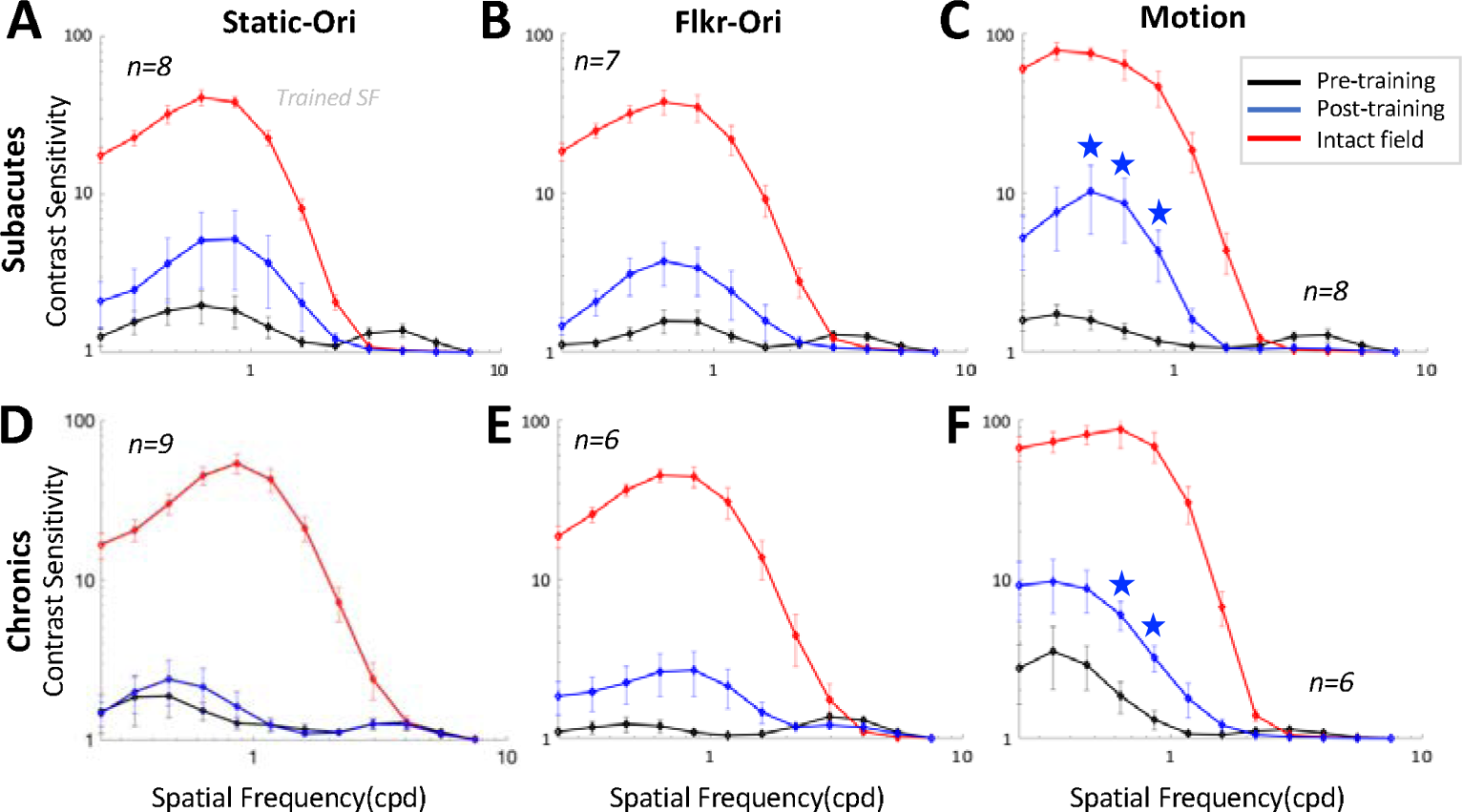
Contrast sensitivity functions (CSFs) for Static-Ori, Flkr-Ori and Motion tasks at selected training locations. **A.** Mean pre-(black line) and post-training (blue line) CSFs for Static-Ori task in subacute participants at selected training locations and corresponding, intact-field locations (red line). Each dot represents averaged CS at a given spatial frequency (mean ± SEM). Grey vertical bar denotes the trained spatial frequency (1cpd). Comparisons at each of the 12 SFs were corrected for multiple comparisons using Bonferroni correction. Bootstrap analysis was performed at each spatial frequency, with blue stars denoting p<0.004. **B.** Average pre- and post-training CSFs for Flkr-Ori in subacute participants at selected training locations and corresponding, intact-field locations. Labeling conventions as in A. **C.** Average pre- and post-training CSFs for Motion in subacute participants at selected training locations and corresponding, intact-field locations. Labeling conventions as in A-B. **D.** Mean pre- and post-training CSFs for Static-Ori in chronic participants at selected training locations and corresponding, intact-field locations. Labeling conventions as in A-C. **E.** Mean pre- and post-training CSFs for Flkr-Ori in chronic participants at selected training locations and corresponding, intact-field locations. Labeling conventions as in A-D. **F.** Mean pre- and post-training CSFs for Motion in chronic participants at selected training locations and corresponding, intact-field locations. Labeling conventions as in A-E.

Bootstrap analysis (**Fig. 8**) revealed a significant change in the qCSFs only for SA and CH participants trained on the Motion task (**Figs. 8C, F**). Both SA (5.7±3.1 post-*vs* 2.3±1.4 pre-training; two-tailed paired Student’s t-test: t_7_=-2.53, p=0.04; **Fig. 8C**) and CH participants (8.4±2.6 post-*vs* 2.6±2.8 pre-training; t_5_=-8.2, p=0.0004; **Fig. 8F**) improved the mean peak CS at Motion-trained locations. Similarly, the mean area under the Motion qCSFs was about 2 times that of pre-training for both SA (3.5±2.8 post-*vs* 1.5±1.3 pre-training; t_7_=-2.82, p=0.03) and CH (4.2±2.1 post-*vs* 1.7±1.8 pre-training; t_5_=-4.68, p=0.005) participants.

Although training was at 1cpd, improved CS for Motion transferred to lower SFs (0.2-0.5cpd– starred in **Fig. 8C, F**) for both SA and CH participants, which was reflected in a reduced, post-training, low-SF truncation level (SA: t_7_=3, p=0.02; CH:t_5_=3.39, p=0.02). However, for both SA and CH participants, post-training AUC and peak CS amplitude at Motion-trained locations remained significantly impaired compared to those at intact-field locations (SA: AUC t_12_=-5.00, p=0.0003; peak ampl. t_8_=-6.81, p=0.0001; CH: AUC t_5_=-4.85, p=0.005; peak ampl. t_5_=-4.79, p=0.005).

## Discussion

The present study examined CS after stroke-induced V1 damage, and asked to what extent this basic property of vision can be restored inside the blind field, whose boundaries were defined using a new, principled approach. Although task performance was sometimes preserved, CS was severely impaired from the earliest time post-stroke, irrespective of whether stimuli were static, flickering, or moving. Repetitive training that exposed contrast-impaired locations to progressively lower luminance contrasts improved CS in most subacute but only half of chronic participants. However, even when improved, CS remained 4-5X lower than normal, suggesting possibly fundamental limitations for recovery of this visual attribute in V1-damaged humans.

### V1 damage impairs CS, even when orientation and direction discrimination are preserved

All CB participants had pronounced deficits in peripheral CS for discriminating static Gabors inside, as well as straddling their perimetrically-defined blind-field border, confirming reports with less precise definitions of the blind field (Das et al., 2014; Saionz et al., 2020) and those using detection rather than discrimination tasks (Ajina et al., 2015; Ajina & Bridge, 2019; Hess & Pointer, 1989; Sahraie et al., 2008; Barbur et al., 1994). Importantly, our earliest (∼2 weeks post-stroke) and chronic participants were similarly affected. This suggests a severe, early deficit in CS for discrimination after V1 stroke in spite of normal CS at intact-field locations, and for many, high performance when discriminating high-contrast stimuli at blind-field locations where CS was poor.

There are mixed reports of preserved *conscious* visual abilities in perimetrically-defined blind fields after V1 damage. For *detection*, some (Ajina et al., 2015; Ajina & Bridge, 2019; Barbur et al., 1994; Blythe et al., 1987; Sahraie et al., 2008), but not others (Hess & Pointer, 1989), reported preservation. For *discrimination*, most failed to find preservation for direction (Azzopardi & Cowey, 2001; Huxlin, 2009; Mazzi et al., 2016; Saionz et al., 2020) or orientation (Das et al., 2014; Mazzi et al., 2016; Morland et al., 1996; Saionz et al., 2020). However, to this point, definitions of the blind-field border have been inconsistent. Here, using a new, principled method for defining this border, we found that ∼half our stroke participants could reliably describe stimuli and perform discrimination tasks at contrast-impaired locations when luminance contrast was 100%. Thus, computations for discriminating large orientation and direction differences inside perimetrically-defined blind fields may be distinct [and longer-lasting post-stroke] than those required for CS. For visually-intact humans, CS for discriminating and detecting low-SF motion stimuli is similar, suggesting that these processes are mediated by the same, directionally-selective mechanisms (Anderson & Burr, 1991; Watson & Robson, 1981; Watson et al., 1980). However, strobe-reared cats, whose early visual neurons fail to develop direction selectivity, display reduced CS for discrimination, but not detection (Pasternak & Leinen, 1986; Pasternak et al., 1985). They can only discriminate direction at high contrasts (Pasternak & Leinen, 1986). Cats with lesions of area 17, the homologue of primate V1, also display greater deficits for discrimination than detection of grating orientation and motion direction (Pasternak et al., 1995). Thus, in V1-damaged humans, residual orientation- and direction-selective units across the residual visual system may support the ability of some individuals to discriminate large orientation or direction differences at high contrast in their blind-fields. However, the loss of a large number of orientation- and direction-selective neurons in V1 means that CS for discriminating these features is severely diminished, even when detection persists.

### Better training efficacy in subacute than chronic stroke participants

Over 90% of subacute but only half of chronic participants exhibited training-induced improvements in CS, despite comparable compliance and number of sessions trained. Additionally, subacutes improved comparably across tasks, whereas chronics improved less on the Static-Ori task. Why should time since stroke make such a difference, especially for static stimuli? A potential factor is progressive, trans-synaptic retrograde degeneration of early visual pathways initiated by the V1 lesion (Beatty et al., 1982; Bridge et al., 2011; Cowey et al., 1989, 2011; Fahrenthold et al., 2021; Herro & Lam, 2015; Jindahra et al., 2009, 2012; Park et al., 2013). Retrograde degeneration is thought to primarily affect parvocellular neurons in the dorsal lateral geniculate nucleus and retina (Cowey et al., 1989) - cells most sensitive to stationary stimuli and low TF content (Derrington & Fuchst, 1979; Foster et al., 1985; Sclar et al., 1990). Subacute participants exhibit fewer signs of retrograde degeneration than chronic participants (Fahrenthold et al., 2021), supporting the notion that better integrity of early visual pathways could underlie their better contrast-training outcomes.

### Training with contrast-varying stimuli does not restore normal CS

When developing therapeutic tools, it is equally important to define functions that can and cannot be restored, and understand why. Although adaptive-contrast training improves CS for direction and orientation discrimination in visually-intact humans (e.g. Levi et al., 2015), we now report its failure to restore normal CS in V1-stroke patients. Instead, it improved CS similarly to training with high-contrast stimuli (Das et al., 2014; Huxlin et al., 2009; Saionz et al., 2020). Given that CS may be strongly related to the activity (Niemeyer & Paradiso, 2017) and number of V1 neurons (Himmelberg et al., 2022; Jigo et al., 2023) representing particular visual field regions, our findings suggest a potentially fundamental limitation of the V1-damaged visual system: with insufficient V1 neurons, the residual circuity may simply be incapable of the processing necessary to restore normal CS over affected parts of the visual field. Thus, in CB participants, the amount of spared V1 representing portions of the blind field (Barbot et al., 2021; Papanikolaou et al., 2014) along with maintenance of its retinal and subcortical input, may be vital to maximize recovery of CS.

### Partial transfer of CS learning to lower spatial frequencies

A central question in perceptual training is whether learning transfers to untrained features. Here, we asked if training with 1cpd contrast-varying Gabors altered the qCSF at untrained SFs. Training with high-contrast, random dot stimuli improves CS for discriminating low-SF drifting gratings in both CB (Das et al., 2014; Huxlin et al., 2009; Saionz et al., 2020) and visually-intact (Levi et al., 2015) humans. Similarly, we saw significant changes in qCSF at low SFs (0.6-1cpd) for motion discrimination in both SA and CH participants. Thus, training with motion stimuli appears to generate improvements in CS, which transfer to lower SFs. After V1 damage, this may be consistent with a greater, relative contribution of spared, motion-sensitive extrastriate areas such as MT/V5, to CS; MT neurons are broadly-tuned but prefer lower SFs (Pawar et al., 2019). In non-human primates, they remain responsive, even months after V1 damage, although direction selectivity decreases over time (Azzopardi et al., 2003; Girard et al., 1992; Hagan et al., 2020; Rodman et al., 1989; Rosa et al., 2000).

### Methodological developments

This study presents a new, principled method for registering behavioral performance measured in-lab with Humphrey perimetry - the most common clinical test for assessing vision loss in CB. On the Humphrey test, when patients cannot see a stimulus 10,000 apostilbs in light intensity, that location is assigned a value of 0dB. Here, we drew the blind-field border as linking sites where sensitivity fell below 10dB on the Humphrey, consistent with the US Social Security Administration definition of blindness. Surprisingly, not only could some CB participants discriminate orientation of high-contrast stimuli fully inside this blind-field border, sometimes, contrast thresholds were severely impaired at locations that straddled the border and included large regions of “intact” vision. We propose that the discrepancy between Humphrey perimetry and in-lab CS measures is rooted in differences between stimuli, tasks, and threshold strategies. Humphrey perimetry displays small (0.43°), broadband (in SF content), white lights on a bright background (10 cd/m^2^) for about 200ms each, whereas our CS tasks used larger, 1cpd Gabors on a brighter background (120 cd/m^2^) for 500ms. Humphrey perimetry also randomly measures detection at equally spaced test locations across the central ∼±21-27° of vision, whereas our discrimination task mapped performance at very few adjacent, overlapping sites, using systematic, 1° lateral displacements of the stimulus after each set of 100-trials. The differences in visual performance resulting from such simple changes in low-level features of stimuli and tasks suggest that great care is needed when defining “blindness” in the context of V1 strokes.

## Conclusions

CS defines thresholds for visibility and discrimination - both fundamental properties of human vision. After a unilateral occipital stroke, CS in the contralateral visual hemifield appears to degrade rapidly, whereas orientation and motion discrimination of high-contrast stimuli can persist for several months. Training protocols that use high-contrast stimuli have been shown to improve detection, discrimination, and identification, but they only partially improve CS at trained, blind-field locations. Here we show that adaptive-contrast training improved CS but failed to restore it to normal levels. However, more subacute than chronic stroke participants benefitted from such training, particularly when discriminating the orientation of static targets. Our results support the notion that CS may be critically dependent on processing within V1. As such, maximizing recovery of CS within cortically-blinded fields may require early intervention and/or preserving the integrity of early visual pathways.

## Data Availability

All data produced in the present study are available upon reasonable request to the authors.

## Acknowledgments

This work was funded by NIH grants R01 EY027314, UL1 TR002001, T32 GM007356, TL1 TR002000, R21 EY031520 and an unrestricted grant to the Department of Ophthalmology at the University of Rochester from Research to Prevent Blindness. The authors wish to thank Christine Callan for patient recruitment, screening and regulatory management. We also wish to thank Rachel Hollar and Terrance Schaefer for performing Humphrey perimetry on all patients.

